# Quantification of positive family history as a risk factor for ischaemic heart disease

**DOI:** 10.1101/2020.02.04.20020560

**Authors:** Basit Iqbal

**Affiliations:** Department of Nuclear Medicine, Gujranwala Institute of Nuclear Medicine & Radiotherapy (GINUM), Gujranwala, Pakistan

**Keywords:** Myocardial ischemia, family history, myocardial perfusion scintigraphy

## Abstract

**Objective:** There is very little research regarding quantification of family history as a risk factor for ischaemic heart disease. This is especially so in the South Asian population, which tends to suffer from ischaemic heart disease at a much younger age, even after environmental and dietary factors are accounted for. This indicates a likely genetic basis. The aim of this study was to quantify family history as a risk factor.

**Methods:** It is a retrospective, cross-sectional study. Patients with family history and hypertension as the only cardiac risk factors were recruited in the study along with control subjects. Myocardial perfusion scintigraphy was used for the detection of myocardial ischemia.

**Results:** 114 patients had hypertension and family history as the only risk factors. 64 of these patients had myocardial ischemia. 102 control patients were also recruited, of whom only 26 suffered from myocardial ischemia. The odds-ratio was thus calculated to be 3.69 (95% CI: 1.54-9.28, p=0.001).

**Conclusions:** These findings suggest a strong genetic basis for ischemic heart disease in the South Asian population. This needs to be taken into consideration when such patients present with non-specific cardiac symptoms.

## INTRODUCTION

Coronary artery disease and subsequent ischaemic heart disease (IHD) have a much higher prevalence amongst the South Asians than other ethnicities, even after all other risk factors are accounted for[1, 2, 3]. Additionally, South Asians tend to have onset of IHD at a much younger age than other ethnicities[2]. Although this may be partially explained by the conventional risk factors, it is likely that genetic susceptibility also has some part to play[3].

Since the Framingham offspring study, numerous studies have suggested that family history (FH) is an independent risk factor for IHD[4, 5, 6, 7, 8, 9, 10]. Some have suggested a genetic basis for it, while others have proposed a predominantly environmental and behavioural basis[11, 12, 13, 14]. However, conflicting data exists regarding the role of FH in providing added value on top of established risk factors in predicting IHD[5, 10]. One of the causes is a lack of a standard definition of FH[8]. Sailam et al have suggested that the Framingham risk score underestimates the FH risk, while Sivapalaratnam, et al have proposed that it fails to provide any useful information[5, 15]. Nasir et al have questioned its extrapolation to non-caucasian population whereas others have questioned its accuracy in women[6, 16].

Most existing studies have used ECG changes, coronary angiography, clinical follow-up, or recently, CT angiography (CTA) as the basis for detection of IHD[6, 8, 10, 17, 18]. The author believes that myocardial perfusion scintigraphy (MPI) is a much more sensitive and specific tool, as it can detect the effects of microvascular abnormalities, and thus needs to be employed as the indicator for myocardial ischemia[18].

Relatively little is known about the significance of familial factors for IHD in the non-caucasian populations, necessitating further studies. In this study, we calculated the significance of FH as a risk factor for IHD in the South Asian population employing MPI as the predictor of IHD.

## METHODS

This is a retrospective, single-centre study. The study was approved by the centre’s institutional review board. The patient population from which the study subjects were included was all the patients presenting for MPI over a period of two years. All patients had presented with non-specific cardiac symptoms such as chest discomfort and/or exertional dyspnoea. No patients with established IHD were included. FH was defined as parent(s) or sibling(s) having onset of IHD before the age of 60 years. As very few patients were referred for MPI with FH as the only risk factor, patients with hypertension (HTN) plus FH as risk factors were selected. Patients with similar age and sex profiles, but only HTN as risk factor were included as controls. This was done so that the odds of suffering from IHD in presence of FH as risk factor could be calculated. The patients were divided into 2 groups, patients with HTN+FH and HTN only group. All patients underwent gated SPECT MPI.

### Statistical Analysis

Statistical analysis was carried out using R: A language and environment for statistical computing by R Foundation for Statistical Computing, Vienna, Austria. Numerical data was reported using mean plus 95% CI. Odds-ratio (OR) for categorical data was calculated using Fisher’s exact test on a 2×2 contingency table. A p-value < 0.05 was considered as statistically significant.

## RESULTS

A total of 1,172 MPIs were carried out during the study period. Patients’ characteristics are shown in table 1. One hundred and fourteen patients fulfilled the inclusion criteria (HTN+FH). Seventy-eight patients (68.4%) were female, whereas 36 patients were male (31.6%). The mean age of the patients was 50.2 years (95% CI: 47.6-52.8). One hundred and six patients presented with chest pain (93%), while 46 suffered from exertional dyspnoea (40%). Sixty-four patients (51%) had ischemia on MPS. One hundred and two age and sex-matched control patients (HTN only) were also included, 64.7% of whom were females and whose average age was 52.8 years (95% CI: 50.1-55.5). The OR of ischemic heart disease in somebody with a positive FH was 3.69 (95% CI: 1.54-9.28, p=0.001).

**Table 1:** Demographic and clinical characteristics of the study patients.

|  | **FH+HTN** | **HTN** |
| --- | --- | --- |
| No. | 114 | 102 |
| Mean age (95% CI) | 50.2 y (47.6-52.8) | 52.8 y (50.1-55.5) |
| Chest pain | 106 | 86 |
| Dyspnoea | 46 | 42 |
| F:M | 78:36 | 66:36 |
| Ischemia | 64 | 26 |

## DISCUSSION

In this study, we were able to quantify the odds of suffering from ischemic heart disease in patients with a positive family history. The number of female patients in the study is larger than males, contrary to the fact that more males suffer from IHD than females, especially in younger age groups. This is because of referral bias, as males frequently tend to have IHD detected by preliminary tests such as ECG and exercise tolerance test, thus resulting in fewer referrals for MPI for detection of inducible ischemia.

The average age of the study subjects was 50.2 years; this is quite in line with published findings[2, 19, 20]. The global average is about 58 years[2]. The higher incidence of heart disease in South Asians has been attributed to dietary and environmental factors[2, 21, 22]. This study strongly suggests that there are genetic factors also at play, since the dietary and environmental factors are accounted for, given that the patients hail from the same environment and have a similar dietary pattern. The predominant complaint of the patients was chest pain followed by dyspnoea. The relative highly odds of 3.69 are similar to published literature[9, 21, 23].

## Conclusion

Cognisant of the fact that this is a retrospective study; the author believes that the findings of this study do suggest a strong genetic basis for IHD in the South Asian population. This needs to be taken into consideration when such patients present with non-specific cardiac symptoms.

The study needs to be expanded upon with a larger cohort and long-term follow up. Similarly, with a larger number of patients, the odds ratio may be calculated separately for the male and female patients.

## Data Availability

Available with author only

